# Falls Efficacy Scale International (FES-I) as a predictor of gait and balance abnormalities in community dwelling older people

**DOI:** 10.1101/2022.09.29.22280485

**Authors:** Lewis McColl, Victoria Strassheim, Matthew Linsley, David Green, Charlotte Dunkel, Heidi Trundle, Jake R Gibbon, Steve W Parry

**Author notes:** **Corresponding Author:** Dr Parry at the Newcastle Hospitals address above.

## Abstract

**Background:** Fear of falling (FoF) is common amongst community-dwelling older adults and is associated with higher falls risk. FoF is common amongst those with gait and balance abnormalities (GABAb), yet the ability of FoF measures to predict GABAb has not been assessed.

**Methods:** Data were reviewed from attendees of the North Tyneside Community Falls Prevention Service. The Falls Efficacy Scale International (FES-I) was used to measure falls efficacy, with a score larger than 23.5 indicating a concern for falling. Gait and balance measures were assessed, with cut-offs used to indicate poor and non-poor results for timed up and go (TUG) (>14s), five times sit to stand (FTSS) (>15s) and gait speed (GS) (<1 m/s). Receiver operating characteristic curves were generated for sensitivity and specificity analysis.

**Results:** FES-I score had good to excellent sensitivity when predicting TUG (87.1%), FTSS (82.9%) and GS results (73.0%) indicative of significant GABAb. Moderate specificity was also observed when predicting GS (62%) and FTSS (62.3%); a low to moderate specificity was observed when predicting TUG (50.0%).

**Conclusion:** A FES-I score of 23.5 or more showed high specificity in identifying those with prolonged TUG and FTSS and slower GS, with moderate specificity.

## Introduction

Approximately a third of community dwelling adults over the age of 65 fall each year (1, 2) with around half experiencing more than one fall per year (3).

Falls are commonly associated with gait and balance abnormalities (GABAb) (4), alongside deficits in strength. Falls not only have the potential to cause serious immediate health issues, such as head injuries and/or fractures, but are also significant source of morbidity and mortality (5), and may lead to long term issues with mobility and a fear of falling (FoF).

FoF is a psychosocial construct encompassing multiple falls related difficulties, including anxiety, loss of confidence and impaired self-efficacy (6). FoF has been reported to have a higher prevalence in women, and increases in incidence with age (7). The measure has also been found to be a predictor of future falls (8, 9), which may be mediated by impairments in physical performance (10). The amelioration of FoF through exercise (11, 12) further suggests a link between FoF and GABAb, themselves a predictor of future falls, mediating the relationship between FoF and falls in community dwelling older adults (13).

Measurement of FoF is commonly done through the Falls Efficacy Scale International version (FES-I) in both clinical and research arenas (14). Our aim was to determine whether FoF, as measured by the FES-I, is associated with GABAb in community dwelling older adults attending a falls prevention service as determined by commonly used gait and balance tests.

## Materials and Methods

### Participants

Routine clinical data (including gait and balance measures, and FES-I scores) was collected from consecutive attendees at North Tyneside Community Falls Prevention Service, full details of which are published elsewhere (15). Caldicott approvals were locally granted, while ethical approval was confirmed as not required by North of England Commissioning Service given the routine anonymised clinical data collection.

### Measures

The FES-I was used as a measure of fear of falling (8), given its reliability, validity, and robustness (16); a score of >23 has previously been used as an indicator of significant concerns around falling (17).

A physiotherapist was responsible for completing a comprehensive physical assessment, including the functional outcome measures below:

1. Gait Speed (GS) as measured by walking speed over a distance of three meters, recorded in seconds. Previous reviews have found a gait speed of 0.8 m/s to be a cut-off in predicting poor clinical outcomes (18).

2. Timed Up and Go (TUG), in which the time taken for a participant to stand from a seated position, walk three metres and return to sit in the chair, is measured in seconds, with a time slower than 14 seconds being considered a predictor of falls (19).

3. The Five Time Sit to Stand (FTSS) test, in which the time taken for a participant to stand from a chair five times without the aid of arms, measured in seconds, with a time slower than 15 seconds being associated with adverse falls and gait and balance issues (20).

### Analyses

Analysis was carried out using SPSS for Windows (Version 25, SPSS, Inc., Chicago, IL, USA). Receiver operating characteristics (ROC) curves were used to estimate the sensitivity and specificity of FES-I scores predicting the presence of a poor outcome for a condition. The area of the graph under the ROC (AUROC) curve was also calculated to provide an estimation of how well the FES-I score discriminated between condition outcomes. Due to the output of the ROC curves within SPSS an FES-I score of 23.5 was used as a cut-off point.

## Results

### Demographics

Nine hundred and ninety-one patients’ data were reviewed (353 males, 638 females). Mean age was 74.5 years (SD= ±8.3 years), mean FES-I score was 28.7 (SD= ±10.2) while 57% reported at least one fall in the year prior.

AUROCs for GS (<1 m/s), TUG (>14s) and FTSS (>15s) are shown in Figure 1, with AUROCs for each measure and their 95% confidence intervals, sensitivity, specificity and positive likelihood ratios provided in Table 1.

**Figure 1:**
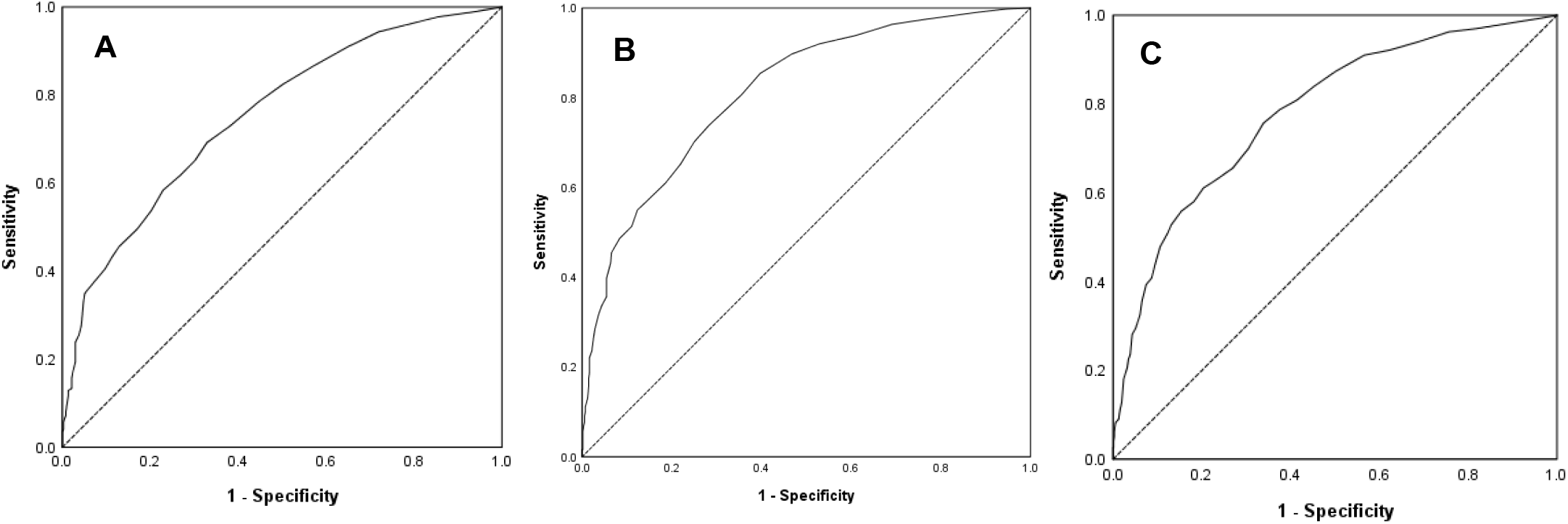
ROC Curves of Physical Function Tests: GS (A), FTSS (B); TUG (C).

**Table 1:** Sensitivity and Specificity analyses generated from ROC curves

| Physical performance measure | AUROC (95% CI) | Sensitivity (%) with FES-I 23.5 | Specificity (%) with FES-I 23.5 | LR+ |
| --- | --- | --- | --- | --- |
| Gait speed (<1.0 m/s) | 0.75 (0.72, 0.78) | 73.0 | 62.0 | 1.92 |
| FTSS (>15s) | 0.80 (0.78, 0.83) | 82.9 | 62.3 | 2.20 |
| TUG (>14s) | 0.78 (0.74, 0.81) | 87.1 | 50.0 | 1.74 |

#### Gait speed

A FES-I score of 23.5 or more had moderate sensitivity (73%) and specificity (62%) in those with a GS of <1 m/s, with higher sensitivity (81.4%) and lower specificity (55.2%) with a slower GS of <0.8 m/s.

#### TUG

A FES-I score of 23.5 or more had high sensitivity (87.1%) with low to moderate specificity (50%) at the >14s cut-off, with a likelihood ratio of 1.74 for a positive test, and 0.26 for a negative test.

#### FTSS

A FES-I score of 23.5 or more had high sensitivity (82.9%) with moderate specificity (62.3%) at the >15s cut-off, with a likelihood ratio of 2.20 for a positive test, and 0.27 for a negative test.

## Discussion

The relationship between falls and FoF is complex, with existing evidence showing that exercise (particularly strength and balance training) ameliorates both (10, 11) Identification and treatment of those with GABAb that predispose to falls is critical in falls prevention. Our study found a FES-I score indicative of FoF was associated with scores on commonly used gait and balance measures that suggest both poor physical function and increased falls risks, with good to excellent sensitivity (73-87.1%), and moderate specificity (50-69.0%). These are broadly similar to sensitivities and specificities for other commonly used clinical tests including faecal occult blood testing and exercising testing in ischaemic heart disease (21, 22).

Our findings are consistent with previous studies that assessed links between FoF and poor physical function, though our comprehensive range of gait and balance measures has never been reported in this context. An association between a slower FTSS and a higher FES-I score has previously been reported (23). The observed association between both a lower GS and TUG, and a FoF was also expected, with slower GS and TUG scores having previously been suggested to either be a deliberate effort to minimise balance difficulties (24), or due to a competition for cognitive processes (25, 26).

Current screening methods for exercise based interventions utilise a multifaceted approach (27), requiring a large number of functional, medical and psychological tests (28), often performed in a secondary care setting. Implementation of a quick, cost-effective and easily administered questionnaire, such as the FES-I, has the potential to allow timely identification of those with GABAb and FoF that may benefit from strength and balance training without the need for trained personnel and clinic time and space, whether primary or secondary care. There is also a current need during the Covid-19 pandemic for screening methods that are not delivered in person; the FES-I may provide just such a proxy measure, though this needs further exploration.

Our findings may provide the initial step in identifying older adults that would benefit from an exercise intervention in a time and resource-effective manner, with a high chance of finding those with GABAb, albeit with a lower ability to distinguish those without. However, the intervention, strength and balance training, is low risk and cost effective, with benefits to all regardless of GABAb, so the risks of harm from FES-I – measured FoF in this context are negligible.

### Strengths and Limitations

The FES-I is an easily implemented and non-labour-intensive method that does not require specialist training to administer or self-complete, making it an ideal tool within clinical practice. Sensitivity was generally high, though specificity was moderate. The study cohort was larger than in similar previous studies (17), however the data were retrospective rather than prospective.

## Conclusion

Fear of falling as assessed by the FES-I is associated with poor performance on commonly used gait and balance measures including the TUG, FTSS and GS. Further work is needed to explore these relationships further, and to assess their potential use in screening for GABAb in the pursuit of improvements in falls prevention.

## Data Availability

All data produced in the present study are available upon reasonable request to the authors

## Acknowledgements

The investigators would like to thank the North Tyneside Community Falls Prevention Service team for their help with data collection and anonymisation.

## Declarations

Ethics approval and consent to participate: All methods were performed in accordance with relevant guidelines and regulations; Caldicott approvals were locally granted, while ethical approval was confirmed as not required by North of England Commissioning Service given the routine anonymised clinical data collection.

Availability of data and materials: The datasets used and analysed during the current study are available from the corresponding author on reasonable request.

Funding: Not Applicable

## Summary

**What is known:** Fear of falling is common amongst community-dwelling older adults and is associated with higher falls risk. FoF is also common amongst those with gait and balance abnormalities.

**What is the question:** Can the FES-I be used to predict poor gait and balance in community dwelling older adults?

**What was found:** FES-I showed high specificity with moderate specificity in identifying poor physical function.

**What is the implication for practice now:** This study shows that further investigation may lead to the potential method for screening falls within the population, potentially reducing falls amongst those with gait and balance disorders. Further prospective analysis is required.

